# Epidemiological Case Study of the 2022 Dengue Outbreak in Makawanpur District, Nepal

**DOI:** 10.1101/2025.04.09.25325521

**Authors:** Rameshwor Parajuli

**Affiliations:** Central Department of Zoology, Tribhuvan University, Nepal

**Keywords:** Dengue, epidemiology, infections, Makawanpur, outbreak

## Abstract

**Introduction:** Makawanpur district, one among 77 districts of Nepal, located in the Bagmati Province, has long been at high risk of contracting dengue. Since 2016 dengue cases were seen in considerable number in the district. Outbreak in 2022 became the worst scenario for this district. The goal of this study is to determine the dengue epidemic scenario in Makawanpur. The study focuses on the dengue outbreak of 2022 at the micro level for the rural municipalities, municipalities and metropolitan city of Makawanpur district.

**Methods:** This study collected the peak epidemic five months of reported dengue cases in 2022 outbreak in Makawanpur from District Health Office (DHO).

**Results:** In 2022 there were 5837 dengue cases reported from Makawanpur which is over 10% of total 54784 cases in that year. In five peak epidemic months of 2022 outbreak in Makawanpur, a total of 5595 cases found with 2514 females and 3081 males. Age-wise dengue infection shows that those between the ages of 20 and 60 are the most affected (3985) and those under the age of five are least affected (176).

**Conclusions:** Geographical location, climatic factors, and socio-economic status of Makawanpur are favorable for prevalence of dengue and Makawanpur can be a potential dengue hub of Nepal.

## 1. Introduction

One of the most common tropical diseases is dengue. About 6.5 million cases and about 7300 deaths from 80 countries worldwide were reported in 2023, the year with the highest number of cases (WHO, n.d.-a). Dengue is still a serious public health issue despite being preventable and curable, especially in Africa and parts of Asia, such as Nepal, where 84% of the population is at risk (WHO, n.d.-b; EDCD, n.d.-a). Out of 6,743 wards in 77 districts, 2,686 wards in Nepal are at risk of dengue transmission; over two lakh individuals reside in high-risk wards, nine lakh in moderate-risk wards, and ten million in low-risk wards (EDCD, n.d.-b).

The virus is mostly carried by female Aedes aegypti and Aedes albopictus mosquitoes. They are typically prevalent in tropical and plain areas. According to Gupta et al. (2018), a recent trend indicates that their breeding is more successful in hilly areas, which may be the reason for the current shift in DENV infection from tropical to temperate regions. While Anopheles mosquitoes breed in clean, stagnant water and are most active at twilight and dawn (Nakase et al., 2023), Aedes mosquitoes breed in stagnant water and are most active in the early morning and late afternoon (Guzman & Harris, 2015). Warm temperatures and high humidity encourage mosquito reproduction (Hales et al., 2002; Murray et al., 2013). Rainy seasons are when dengue and malaria are most common (Gubler, 1998; Descloux et al., 2012). Anopheles mosquitoes grow best in areas with big bodies of water or wetlands (Messina et al., 2019).

The first dengue case in Nepal was reported in 2004 based on genetic similarities. It is believed that dengue originated in India. There are five serotypes of the dengue virus (DENV1–5) that can infect humans. In Nepal, four dengue virus serotypes were discovered in 2006 (Rijal et al., 2021). Out of the four serotypes identified in the 2006 outbreak, DENV1 was more common in the 2010 and 2016 outbreaks, while DENV2 was more common in the 2013 and 2019 outbreaks. The epidemiological trajectory of dengue epidemic reveals 3-year interval outbreak (2010-2013-2016-2019 and now 2022) (Adhikari & Subedi, 2020).

Up until 2010, dengue was mostly found in Nepal’s terai, but in 2016, it started to spread to the country’s mountainous areas. Weak diagnostic facilities, little vector research, unplanned housing, congested populations, and poor waste management are the most likely causes of the rise in dengue incidence during the past five years. Tools for DENV control include better basic research and epidemiological training programs for local scientists and lab staff (Neupane et al., 2014).

## 2. Materials and Methods

### 2.1. Study design

This is a study analyzing the reported data from District Health Office (DHO), Makawanpur. This study’s data depicts a serological diagnosis made with an IgM ELISA and a fast test kit. The data in this study covers a period of five epidemic months (July-November) of 2022 outbreak in Makawanpur.

### 2.2. Study Area

Makawanpur is a district located in Bagmati province of Nepal. It is one among 77 districts of Nepal and one among 13 districts of Bagmati province. There are seven provinces in Nepal. Makawanpur district is bordered north by Kathmandu and Dhading districts, to the west by Chitwan district, to the east by Lalitpur and to the south by Parsa. Makawanpur consists of 10 Municipalities, out of which one is a sub-metropolitan city (Hetauda), one is an urban municipality (Thaha) and eight are rural municipalities.

Figure 1. below shows the study area in national and provincial map.

**Figure 1.**
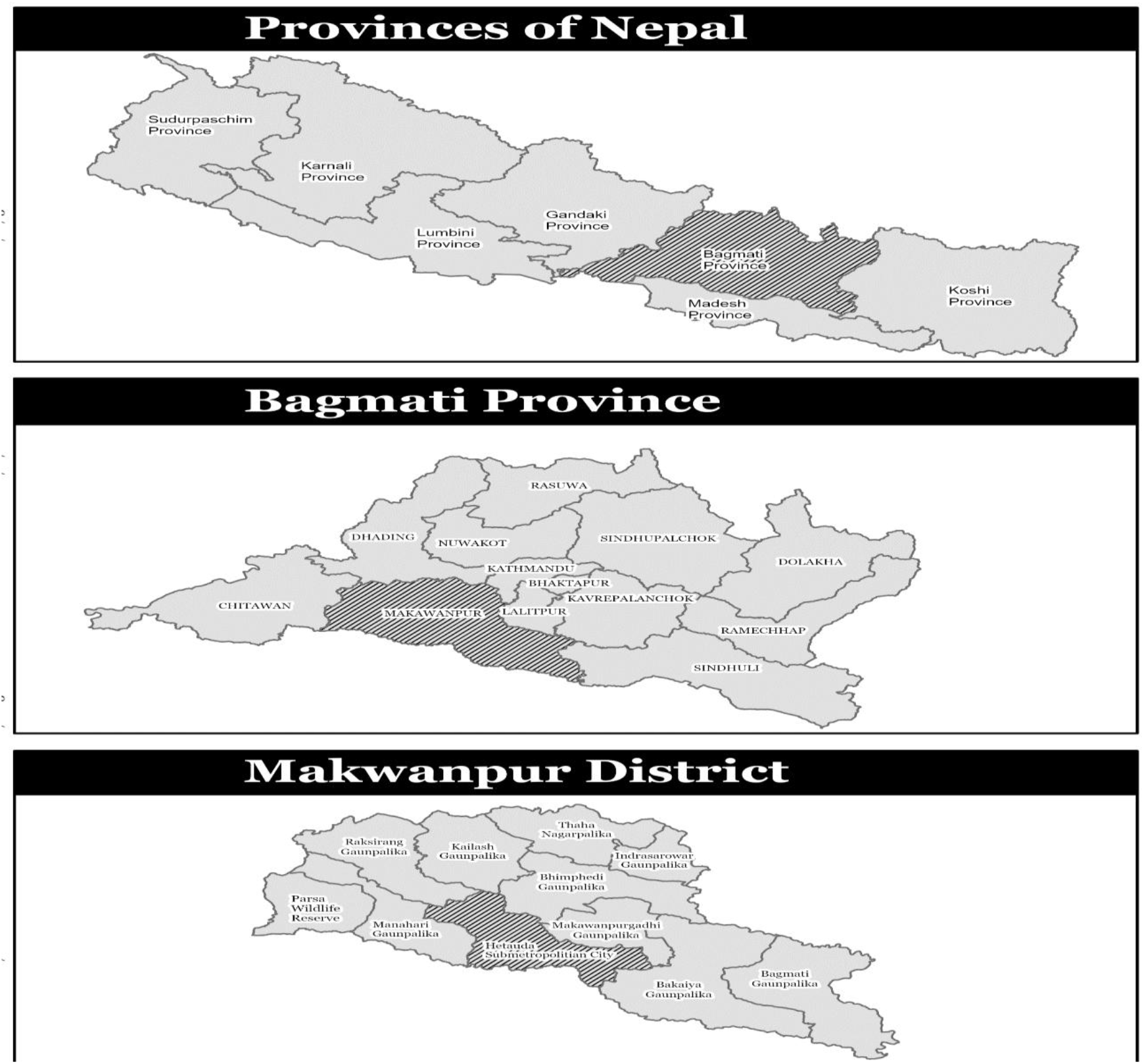
Study area in provincial and national map.

### 2.3. Data Collection

Dengue data of peak epidemic five months (July-November) in Makawanpur was collected from the records of laboratories, health centers, hospitals, and medical facilities.

### 2.4. Data Analysis

Data were first entered and properly arranged in Microsoft Excel 2010 to perform the analysis.

### 2.5. Study Approval

Central Department of Zoology - Tribhuvan University (CDZ-TU), Nepal recommended this study and on the basis of this recommendation, DHO Makawanpur approved this study.

## 3. Results

### 3.1. Makawanpur becoming a potential hub of dengue in future

Makawanpur’s share of total dengue cases varied widely from only 0.14% in 2017 to over 10% in 2022. The year 2022 saw a massive rise in Makawanpur with 5837 cases and 3 deaths accounting for 1 in 10 cases nationwide.

Figure 2 below visualizes a clear peaking of dengue in Makawanpur district

**Figure 2.**
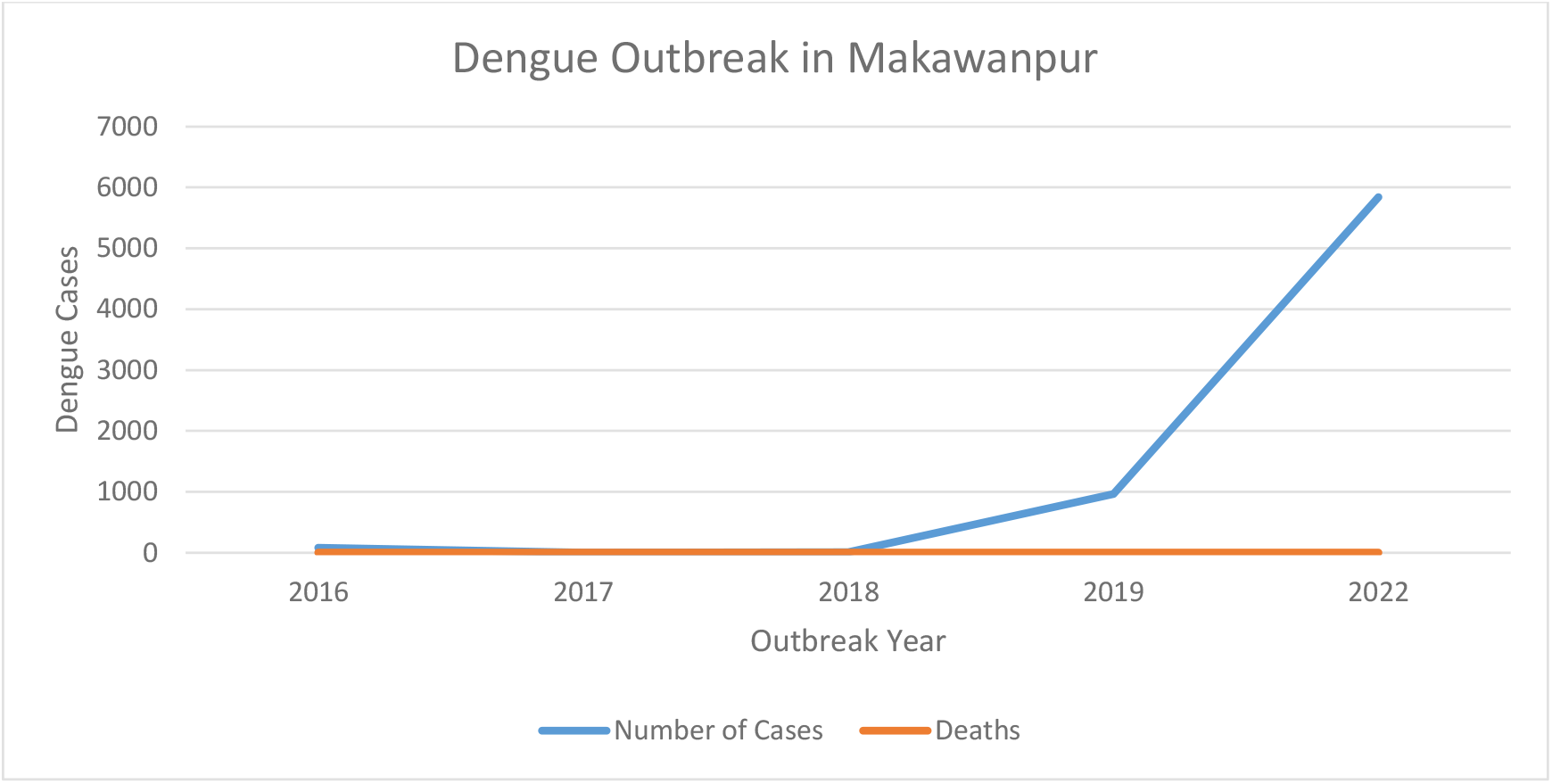
Dengue in Makawanpur from 2016-2022.

### 3.2. Dengue in local level of Makawanpur in 2022 outbreak

Out of total 5595 cases reported in five peak epidemic months from DHO Makawanpur; Indrasarowar Rural Municipality reported 4 cases, Kailash Rural Municipality (6 cases), Bagmati Rural Municipality (8 cases), Raksirang Rural Municipality (24 cases), Bakaiya Rural Municipality (43 cases), Bhimphedi Rural Municipality (55 cases), Makwanpurgadhi Rural Municipality (75 cases), Thaha Municipality (173 cases), Manhari Rural Municipality (932 cases), and Hetauda Sub-Metropolitan City reported the highest 4269 cases. Table 1 below presents the distribution of cases in the local level.

**Table 1.** Distribution of Cases by Local Level in Makawanpur in 2022 outbreak.

| Local Level | No. of Cases | % of Total Cases (out of 5595) |
| --- | --- | --- |
| Indrasarowar Rural Municipality | 4 | 0.07% |
| Kailash Rural Municipality | 6 | 0.11% |
| Bagmati Rural Municipality | 8 | 0.14% |
| Raksirang Rural Municipality | 24 | 0.43% |
| Bakaiya Rural Municipality | 43 | 0.77% |
| Bhimphedi Rural Municipality | 55 | 0.98% |
| Makwanpurgadhi Rural Municipality | 75 | 1.34% |
| Thaha Municipality | 173 | 3.09% |
| Manhari Rural Municipality | 932 | 16.67% |
| Hetauda Sub-Metropolitan City | 4269 | 76.40% |
| Total | 5595 | 100% |

### 3.3. Dengue is more prevalent among active age group males in Makawanpur

During the five months of the outbreak in 2022, out of total 5595 cases; 2514 were females (44.93%) and 3081 were males (55.07%) affected. Looking at the age-wise data, those between the ages of 20 and 60 were the most affected (3985), followed by those between the ages of 6 and 19 (1045), those over 60 (363), and those under the age of five (176) were least affected. Table 2 below shows the population affected by dengue in Makawanpur on the basis of age group in 2022 outbreak.

**Table 2.** Population affected by dengue on the basis of age group in Makawanpur in 2022 outbreak.

| Age Group (Years) | Number of Cases | Remarks |
| --- | --- | --- |
| Under 5 | 176 | Least affected |
| 6–19 | 1,045 | Moderately affected |
| 20–60 | 3,985 | Most affected |
| Above 60 | 363 | Less affected |

## 4. Discussion

### 4.1. Overall dengue trend in Makawanpur

DENV cases has been rising rapidly since the first case seen in 2004 in Japanese traveler who came through India. The first outbreak just happened after 2 years in 2006 where 32 cases reported, and this outbreak occurred in Chitwan district which is immediate neighboring district of Makawanpur (Adhikari & Subedi, 2020). Another outbreak occurred in 2010, then in 2013, 2016 and 2019. This shows 3-year interval outbreak happening since 2010. Though no cases were reported from Makawanpur in 2006 but still there is high chance that many cases might have left undiagnosed or unreported.

Onward 2016 considerable dengue effects are being reported from Makawanpur; 82 cases were reported in 2016; among the three deaths that happened in 2017 due to dengue virus infection, one was reported from Makawanpur district and remaining two from Chitwan and Palpa (Rijal et al., 2021); among the three deaths happened in 2018 due to the dengue virus infection, one was reported from Makawanpur district and remaining two deaths reported from Rupandehi. Major devastative situation in Makawanpur occurred in 2019 and 2022 outbreaks. In 2019, 961 cases were reported and in 2022 outbreak, 5837 cases were reported. The six districts in the centrally located province of Bagmati (Kathmandu, Lalitpur, Bhaktapur, Kavrepalanchowk, Makwanpur, and Dhading) exhibited statistically higher dengue incidences in 2022 outbreak (Bhandari et al., 2025).

If we see the overall trend from 2006 – 2019, 85% cases are reported from three district; Chitwan, Parsa and Jhapa (Adhikari, 2020). Makawanpur district was fourth highest dengue reporting district in 2022 AD outbreak (EDCD, n.d.-c). We cannot ignore the fact that Makawanpur is located in between Parsa and Chitwan. A retrospective analysis of dengue outbreak fatalities in 2022 in Nepal shows that out of 88 deaths from all 77 districts, the majority of the deaths were reported from districts of Bagmati province (60.2%, 53 deaths) where Makwanpur district reported 3 deaths following Chitwan (4), Lalitpur (11) and Kathmandu (23) respectively (Kandel et al., 2025).

This study shows that those areas connected directly to highway and densely populated has a greater number of dengue cases; for instances Hetauda, Thaha and Manhari (Table 1). This is consistent with the findings of a case study in Nepal evaluating the dengue outbreaks in a fragile state (Griffiths et al., 2013). Also, our study shows that the male is more infected than female and this finding is consistent with a study done in Nepal assessing seven years of dengue emergence surrounding 2010 epidemic, which finds that male is more exposed to the vectors causing dengue fever than female because male is working and earning member of a family in a typical Nepalese society (Pandey et al., 2015).

A study of dengue virus surveillance in Nepal finds that male is more infected than female from dengue virus at the ratio of 1.17:2.5 and 15-40 years’ age group population are most vulnerable whereas 40+ age group are less infected than other age group population (Gupta et al., 2018). It is consistent with the data from 2022 outbreak in Makawanpur showing that the highest affected number of population is 20 to 60 years’ age group (3985 cases out of 5595) and 60+ age group least affected with just 363 cases (Table 2). Another study in southern Vietnam finds that active age group are more exposed to the vectors and hence more susceptible to dengue disease (Thai et al., 2011).

### 4.2. Climatic factors, geographical location and rapid urbanization of Makawanpur

Dengue outbreaks have increasingly spread from plain tropical lands to the hills and mountains in Nepal. Temperature and relative humidity are the major contributing factors in dengue epidemics (Tuladhar et al., 2019). Makawanpur district features diverse topography ranging from the flatlands of the Terai to rolling hills and forested highlands. Elevation varies from place to place, from around 166 meters in the lowlands; Hetauda sub-metropolitan city to 2,588 meters at Daman; Thaha municipality and the district has a subtropical to temperate climate (Nepal Database, 2025).

A study regarding temperature-driven dengue transmission changes over the worldwide shows that countries experiencing large increases in dengue incidence were generally those in subtropical regions at high latitudes, with the most notable changes occurring in South Asia (Feng et al., 2024). Furthermore, another study done in a range of altitude to find dengue prevalence shows that below 500-meter altitude has greatest risk of dengue, 500-1500m has moderate risk and above 1500m has the lowest risk of dengue virus infection (Gyawali et al., 2020).

In addition, Makawanpur district is the transit hub of Nepal and its Headquarter is Hetauda sub-metropolitan city, the capital city of Bagmati province. Hetauda is surrounded with the major cities of Nepal like: the capital city, Kathmandu, 80 km away in north; largest border of Nepal and India, Birgunj (Parsa District), just 55 km away in south; and a massively growing commercial city, Narayanghar (Chitwan District), 75 km in west. Our study finds that among all regions of Makawanpur district, Hetauda city reported the 76.3 % of dengue fever (4269 out of 5595). A study done in Medellin Colombia shows that the zones with high public transportation and human movement have more dengue prevalence and are the major drivers of dengue incidence and dengue dynamics (Shragai et al., 2022).

Hetauda is rapidly undergoing urbanization in recent years. Hetauda became capital city of Bagmati province in 2020 AD, which is five years after the constitution of Nepal, 2015 got declared. Due to this, government agencies, government bodies, private agencies working in partnership with government and related institutions either shifted to or originated in Hetauda. Private companies, industries, and housing sectors rapidly relocated to Hetauda and its periphery. Many earlier studies shows that rapid urbanization and inadequate infrastructure (e.g., poor waste management, water storage practices) are associated with dengue outbreaks (Kroeger et al., 2006; Focks, 2003). Studies also shows that increased travel and trade is positively associated with the frequency and intensity of dengue outbreaks (Wilder-Smith & Gubler, 2008a). Public movement from endemic to non-endemic region can introduce parasites and viruses into the new region and facilitate the dengue transmission (Wilder-Smith & Gubler, 2008b; Harish et al., 2024).

### 4.3. Socio-economic and demographic factors of Makawanpur

Chepangs are the ethnic groups of Nepal classified as highly marginalized and has no representation in any socio-political spheres on the basis of socio-economic indicators (Indigenous Voice, n.d.). Although the Chepang settlement is currently dispersed throughout half of the country, 37% of them reside in Makwanpur (Byanjankar, 2024). Another major ethnic group called Tamang, often referred to as “Janajati”, are marginalized group of Nepal and are third largest indigenous group in Nepal. Makawanpur district has nearly 50% Tamang ethnic group (National Population and Housing Census [NPHC], 2021). When compared to other ethnic groups in Nepal, the Tamang have the highest poverty rate, at 28.34% and their per capita income is also low NPRs. 33541 falling below the national average (Sapkota, 2023).

Both Chepang and Tamang are lower educated ethnic groups and have no good access to information and technology. A structured questionnaire done in five districts of Nepal showed that people with low socio-economic status and literacy had low level of knowledge, fair practice and satisfactory attitude towards dengue (Dhimal et al., 2014). A study done on the influence of socio-economic, demographic factors on the regional distribution of dengue in the United States and Mexico draws three main lessons for increase in dengue incidence: first; low economic status is generally associated with a rise in the incidence of dengue, second; a more granular analysis revealed that the crucial factor is less education among people and third; lack of access to information or technological infrastructure (Watts et al., 2020). Similar study from Nepal on socio-economic and demographic determinants of Dengue Hemorrhagic Fever (DHF) reveals that in order to effectively manage and control dengue outbreaks, targeted interventions based on thorough socioeconomic analyses are crucial and dengue incidence throughout Nepal are influenced by socioeconomic factors (Rk & Km, 2025).

There are a few limitations to this study. The majority of dengue cases in our study were diagnosed using ELISA (IgM/IgG antibody detection), which may have failed to identify dengue viral infection in its early stages and may have produced false-positive results in patients who had previously contracted dengue or any other flavivirus infection. In Nepal, systematic specific diagnostic technologies like polymerase chain reaction (PCR) for viral genome identification and viral isolation are not in practice. In addition, serological instruments and examinations employed even in the midst of epidemic outbreaks are not the gold standard, and it is possible that the DENV virus would go undetected before antibodies emerge, which would significantly restrict precise diagnosis during epidemics (Gupta et al., 2018).

## 5. Conclusions

Makawanpur has remained a susceptible region to dengue virus disease since 2016. A sharp increment in dengue cases in 2019 and 2022 outbreaks show that the prevalence of dengue virus in Makawanpur is rapidly rising in recent years and may experience further inclination. The variables like geographical location, climatic factors, socio-economic status, demographic characteristics, and rapid urbanization are likely to promote dengue prevalence in Makawanpur. Neighboring districts of Makawanpur with high dengue infections can play a role in transmitting the dengue. Active age group (20-60 years) males need to follow more precautions to be safe from dengue infection. Timely measures like proper housing management and waste management, awareness and education among people, good surveillance and accurate diagnostics are crucial to control dengue before it’s too late.

## Data Availability

All data produced in the present study are available upon reasonable request to the author.

## Acknowledgments

I thank the Health Office Makawanpur and all the in-charge of respective labs, hospitals, and health centers. I would like to thank the Central Department of Zoology, Tribhuvan University for their co-ordination in this study.

## Competing interests statement

The author has no competing interests to declare.

## Data Availability Statement

Data supporting this study can be available upon a reasonable request to the corresponding author.

## Ethics Statement

No humans and animals were touched or harmed in the study. Approval for the study was taken from the government authority; DHO Makawanpur (Ref No: 079-080) and institutional recommendation; CDZ-TU, was obtained prior to this study.

## Funding

This research received no specific grant from any funding agency in the public, commercial or not-for-profit sectors.

## Notes

### Competing Interest Statement

The authors have declared no competing interest.

### Funding Statement

This study did not receive any funding.

### Author Declarations

The Central Department of Zoology, Tribhuvan University, recommended that the Health Office of Makawanpur district grant access to dengue data. DHO Makawanpur accepts the recommendation and grants access to the data, which are line-listed and available upon a genuine and reliable request. The datasets used in this study were individual-level data and had been de-identified before their use in this study.

### Summary of Updates

This updated version has a more descriptive nature and includes much more depth in the discussion.

